# Impact of dopamine deficiency and REM sleep behavior disorder on cognition in early neuronal synuclein disease with hyposmia

**DOI:** 10.1101/2024.12.12.24318917

**Authors:** Daniel Weintraub, Anuprita R Nair, Ryan Kurth, Michael C. Brumm, Michele K. York, Roseanne Dobkin, Kenneth Marek, Caroline Tanner, Tanya Simuni, Andrew Siderowf, Douglas Galasko, Lana M. Chahine, Christopher Coffey, Kalpana Merchant, Kathleen L. Poston, Tatiana Foroud, Brit Mollenhauer, Ethan G. Brown, Karl Kieburtz, Mark Frasier, Todd Sherer, Sohini Chowdhury, Roy N. Alcalay, Aleksandar Videnovic, the Parkinson’s Progression Markers Initiative

## Abstract

**Objectives:** To determine the impact of dopamine deficiency and isolated REM sleep behavior disorder (iRBD) on cognitive performance in early neuronal alpha-synuclein disease (NSD) with hyposmia.

**Methods:** Using Parkinson’s Progression Markers Initiative baseline data, cognitive performance was assessed with a cognitive summary score (CSS) developed by applying regression-based internal norms derived from a robust healthy control (HC) group. Performance was examined for participants with hyposmia classified as NSD-Integrated Staging System (NSD-ISS) Stage 2, either Stage 2A (CSF alpha-synuclein seed amplification assay [SAA]+, SPECT dopamine transporter scan [DaTscan]-) or 2B (SAA+, DaTscan+).

**Results:** Participants were Stage 2A (N=101), Stage 2B (N=227) and HCs (N=158). Although Stage 2 overall had intact Montreal Cognitive Assessment scores (mean (SD) =27.0 (2.3)), Stage 2A had a numerically worse CSS (z-score mean difference =0.05, p-value NS; effect size=0.09) and Stage 2B had a statistically worse CSS (z-score mean difference =0.23, p-value <0.05; effect size=0.40) compared with HCs. In Stage 2A participants with hyposmia alone had normal cognition, but presence of comorbid iRBD was associated with significantly worse cognition (z-score mean difference =0.33, p-value <0.05, effect size =0.50). In Stage 2B participants with hyposmia had abnormal cognition (z-score mean difference =0.18, p-value =.0078, effect size =0.29), and superimposed iRBD had a non-statistically significant additive effect.

**Interpretation:** Using a CSS, early NSD with hyposmia is associated with measurable cognitive deficits compared with robust HCs, particularly in presence of dopamine system impairment or comorbid iRBD, highlighting the importance of focusing on cognition in early-stage synuclein disease.

## INTRODUCTION

Long-term significant cognitive impairment is a common and feared outcome in Parkinson’s disease (PD), with dementia affecting up to 80% of patients in the long-term(1, 2). Mild cognitive impairment (MCI) is also relatively common(3), even in *de novo* or early disease(4–6). In addition, subtle cognitive changes can be detected in the key prodromal states, including in persons with hyposmia(7), a prodromal symptom for PD, or in patients with isolated REM sleep behavior disorder (iRBD)(8), a prodromal disorder for both PD and dementia with Lewy bodies (DLB). One study found both global and specific cognitive deficits in persons with both hyposmia and iRBD(9).

Recently we proposed a research biological definition and staging system for neuronal α-synuclein disease (NSD-ISS)(10), with NSD defined by the presence of biomarker of pathological neuronal α-synuclein species as the primary biological anchor, and dopaminergic neuronal dysfunction as a subsequent additional biological anchor. NSD also incorporates an integrated staging system (ISS), rooted in the aforementioned biological anchors plus severity of functional impairment caused by clinical signs or symptoms along three tracks (i.e., motor, non-motor and cognitive). The presence of clinical signs marks the transition to NSD-ISS stage 2 and beyond, with Stage 2 characterized by *subtle* signs or symptoms (e.g., hyposmia or iRBD, or early motor or cognitive symptoms) but without functional impairment (i.e., prodromal disease). Stage 2A is neuronal α-synuclein positive but without dopaminergic neuronal dysfunction (negative), while Stage 2B is neuronal α-synuclein positive and with dopaminergic neuronal dysfunction present (positive). In Parkinson’s Progression Markers Initiative (PPMI) the presence of neuronal α-synuclein is currently determined by a positive cerebrospinal fluid (CSF) α-synuclein seed amplification assay (SAA), and dopaminergic neuronal dysfunction by a single-photon emission computerized tomography (SPECT) dopamine transporter (DAT) scan (DaTscan).

When assessing cognitive abilities, an option that combines the richness of a neuropsychological battery with the simplicity of a single test score is a cognitive summary score (CSS). In the Parkinson Associated Risk Syndrome (PARS) study, a CSS was used to document subtle cognitive changes in prodromal PD (i.e., persons with hyposmia and positive DAT scan)(7, 11). Using the original PPMI cognitive battery (five detailed, well-established cognitive tests)(12), we recently implemented a robust norming process using baseline PPMI data to enhance sensitivity in detecting cognitive changes in *de novo* PD compared with healthy controls (HCs), in a fashion similar to that employed to update the norms for the Dementia Rating Scale(13).

The goals for these analyses were to use the proposed NSD-ISS criteria to assess cognition in persons with early-stage NSD (i.e., Stage 2), on the basis of having (1) a positive CSF neuronal α-synuclein SAA test, with and without dopaminergic neuronal dysfunction, and (2) hyposmia. We hypothesized that persons with early-stage NSD and hyposmia would have worse cognitive performance compared with HCs, and that both co-morbid iRBD and dopaminergic neuronal dysfunction would increase the severity of cognitive deficits. Establishing this would lend support to the current inclusion of a cognitive track in NSD-ISS, from the earliest disease stages, and more importantly highlight the importance of including cognition as an outcome measure in disease-targeting randomized controlled trials for early stage NSD.

## METHODS

We adhered to the STROBE checklist for cross-sectional studies(14).

### Enrollment Cohorts

The PPMI study and enrollment cohorts, all of whom are represented in these analyses, have been extensively described(15, 16). All participants signed an approved informed consent form. Individuals without a known diagnosis of PD are enrolled in the study based on risk factors for or signs/symptoms of potential early synucleinopathy (also called prodromal disease), including hyposmia, iRBD and relevant genetic variants carriers as described previously(17). Inclusion criteria for *de novo* PD participants included: (1) an asymmetric resting tremor or asymmetric bradykinesia, or at least two symptoms out of resting tremor, bradykinesia, and rigidity; (2) a recent clinical diagnosis of PD (mean [SD] duration from diagnosis = 8.5 [7.3] months); (3) being untreated with PD medications at study entry; (4) evidence of dopaminergic neuronal dysfunction based on DAT SPECT imaging; and (5) being non-demented by site investigator clinical determination. Participants with DAT scans without evidence for dopaminergic deficit (SWEDDs) have also been previously described(16).

HCs at enrollment were required to (1) not have clinically significant neurological dysfunction, (2) not have a first-degree relative with PD, and (3) have a Montreal Cognitive Assessment (MoCA)(18) score ≥27. For the purposes of these analyses HCs could not have hyposmia and had to have a negative CSF neuronal α-synuclein SAA test. Finally, to generate the internally-derived norms for the CSS, we created a robust HC subgroup that was also required to have a year 1 MoCA score of ≥26 and not have had more than a 2-point drop in their MoCA score between baseline and year 1. This group is hereafter called the HC subgroup.

### Redefining *de novo* PD participants using NSD-ISS

All the aforementioned enrollment cohorts were reclassified to their NSD-ISS stage. CSF was collected as previously described and assayed for neuronal alpha-synuclein with seed amplification assay (SAA) test as described(19). Dopaminergic dysfunction was assessed with dopamine transporter SPECT (DaTscan) as described(16).

Only participants classified as Stage 2 (i.e., those with *subtle* signs or symptoms [i.e., no functional impairment], also called prodromal disease) were included in these analyses. Thus, participants classified as Stage 3 (slight signs or symptoms [i.e., slight functional impairment])(17) and above were excluded from these analyses.

Participants with normal and abnormal DaTscan results were classified as Stages 2A and 2B, respectively. For the purposes of these analyses a DaTscan deficit was defined as <75% age- and sex-expected lowest putamen specific binding ratio. In addition, the following inclusion criteria were also applied: participants were required to: (a) have hyposmia, defined by an age/sex-adjusted University of Pennsylvania Smell Identification Test (UPSIT) ≤15^th^ percentile (those who were normosmic or had a missing UPSIT were excluded); (b) not be on medication for treating the symptoms of PD; (c) have completed all five tests in the original PPMI cognitive battery; and (d) not have cognitive, motor, or other non-motor features qualifying for Stage 3 or higher. Lastly, participants were classified into “hyposmia only” and comorbid “hyposmia and iRBD” subgroups. To make these subgroups as “pure” as possible, only participants *without* any evidence of possible RBD (defined by a score ≥ 6 on the RBDSQ, self-reporting an RBD diagnosis without PSG confirmation, or self-reporting dream enactment behavior) were included in the hyposmia group, and only participants *with* polysomnogram (PSG)-confirmed RBD were included in the iRBD group. NSD participants with a normal DaTscan were stage 2A, and those with an abnormal DaTscan were stage 2B.

Mapping the biologically-defined NSD-ISS stages to the three primary PPMI enrollment cohorts (i.e., de novo PD, prodromal PD and healthy controls), all HCs were enrolled as HCs, about half of Stage 2A participants were enrolled as hyposmics and most of the rest qualified on the basis of having iRBD (and also were subsequently determined to have comorbid hyposmia), and about half of the Stage 2B participants were enrolled as sporadic PD, with nearly all the rest enrolled as hyposmic or having iRBD.

### Cognitive summary score

Baseline data were used, at which point those participants enrolled in the PD cohort were recently diagnosed and had not yet begun treatment with PD medication. The following steps were taken: (1) creating a robust HC subgroup that did not demonstrate cognitive decline over time; (2) using the HC subgroup to create regression-based internally-derived standardized scores (z-scores) for six cognitive scores across five tests, with the standardized scores controlling age, sex and education; and (3) creating a CSS by averaging all standardized test z-scores. The original cognitive battery was used to create the CSS to optimize the amount of data available for analyses. This original battery was composed of the Hopkins Verbal Learning Test-Revised (HLVT-R immediate and delayed free recall scores)(20), the Benton Judgment of Line Orientation - 15 item version (JLO)(21), Symbol-Digit Modalities Test (SDMT)(22), Letter-Number Sequencing (LNS)(23), and semantic (animal) fluency(24). Together these tests assess memory, visuospatial function, information processing speed, executive function, working memory and language. Global cognition was assessed with the Montreal Cognitive Assessment (MoCA)(18).

### Data availability

Data used in the preparation of this article were obtained on February 05, 2024, from the PPMI database (www.ppmi-info.org/access-data-specimens/download-data), RRID:SCR_006431. For up-to-date information on the study, visit www.ppmi-info.org. The analyses were conducted by the PPMI Statistics Core and used actual dates of activity for participants, a restricted data element not available to public users of PPMI data. The PPMI Data Access Committee approved use of the α-syn SAA and DaTscan results for participants. Statistical analysis codes used to perform the analyses in this article are shared on Zenodo [10.5281/zenodo.14047599]. Information about the PPMI protocol can be found on protocols.io: https://dx.doi.org/10.17504/protocols.io.n92ldmw6ol5b/v2

### Analyses

Statistical analyses were performed using SAS v9.4 (SAS Institute Inc., Cary, NC; sas.com; RRID:SCR_008567). Using PPMI baseline data participant characteristics were compared between the Stage 2A, Stage 2B and HC groups using mean (standard deviation) and median (interquartile range) for continuous measures and frequency (percentage) for categorical measures. A t-test was performed for continuous measures and a Chi-square test was performed for categorical measures to compare characteristics across the three groups. Next, a t test was employed to assess p values for difference in mean MoCA score between Stage 2A and Stage 2B groups. Cohen’s d effect sizes were also evaluated and reported.

To evaluate the impact of superimposed iRBD (in addition to hyposmia) on cognitive performance in Stage 2 participants, two-sample t tests were performed to compare the CSS between hyposmia only vs. comorbid hyposmia and iRBD participants. These subgroups were also compared to the HC subgroup. The associated Cohen’s d effect sizes were also evaluated and reported. This analysis was performed for the overall Stage 2 group, as well as based on their dopaminergic dysfunction status (i.e., separately within Stage 2A and Stage 2B subgroups).

## RESULTS

### Cohorts

The cohort comprised of NSD Stage 2 participants (2A N=101, 2B N=227) and HCs (N=158). Of the 328 participants with Stage 2 NSD and hyposmia, 27.7% (N=91) had comorbid iRBD, representing 43% of the Stage 2A cohort and 21% of the Stage 2B cohort.

### Demographic and clinical characteristics

Regarding cohort characteristics, 60-65% of all three cohorts were male, consistent with PD research cohorts (**Table 1**). HCs were significantly younger than Stage 2 participants. All Stage 2 participants met non-motor criteria for Stage 2, as an inclusion criterion for these analyses was presence of hyposmia. Participants can also meet Stage 2 criteria on the basis of subtle cognitive changes (i.e., MDS-UPDRS Part 1 cognition item = 1 and MoCA score ≥25), and 17% of Stage 2A and 16% of Stage 2B participants met cognition criteria for Stage 2, compared with 9% of HCs. Close to 60% of Stage 2B participants met the motor criteria for Stage 2. 38.5% (35/91) of participants with co-morbid iRBD were taking clonazepam as symptomatic therapy, compared with <1% of hyposmia alone participants (1/237).

**Table 1.**
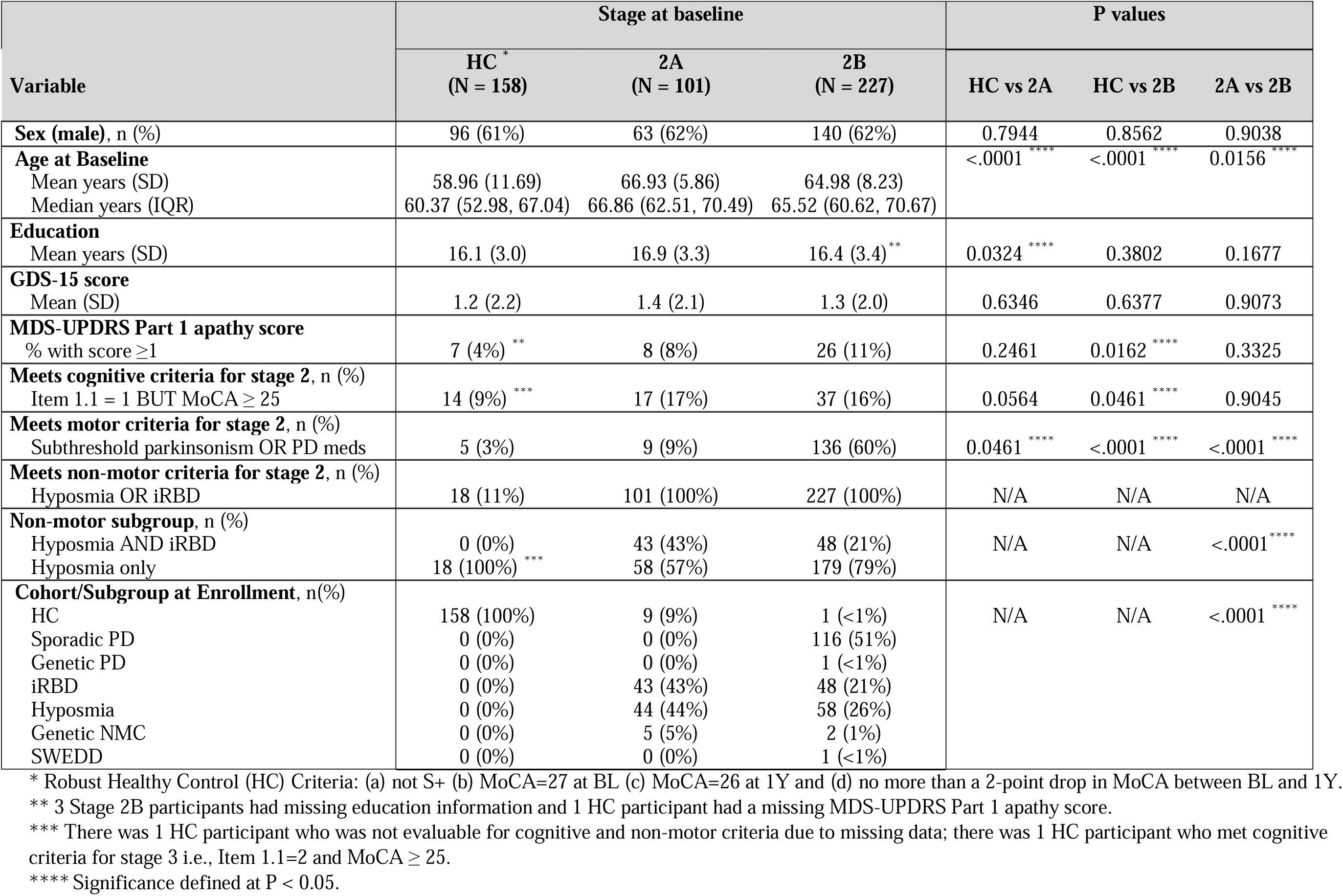
Characteristics of NSD Stage 2 and HC subgroup participants.

| Variable | Stage at baseline |  |  | P values |  |  |
| --- | --- | --- | --- | --- | --- | --- |
|  | HC<sup>*</sup><br>(N = 158) | 2A<br>(N = 101) | 2B<br>(N = 227) | HC vs 2A | HC vs 2B | 2A vs 2B |
| **Sex (male), n (%)** | 96 (61%) | 63 (62%) | 140 (62%) | 0.7944 | 0.8562 | 0.9038 |
| **Age at Baseline** |  |  |  | <.0001<sup>****</sup> | <.0001<sup>****</sup> | 0.0156<sup>****</sup> |
| Mean years (SD) | 58.96 (11.69) | 66.93 (5.86) | 64.98 (8.23) |  |  |  |
| Median years (IQR) | 60.37 (52.98, 67.04) | 66.86 (62.51, 70.49) | 65.52 (60.62, 70.67) |  |  |  |
| **Education** |  |  |  |  |  |  |
| Mean years (SD) | 16.1 (3.0) | 16.9 (3.3) | 16.4 (3.4)<sup>**</sup> | 0.0324<sup>****</sup> | 0.3802 | 0.1677 |
| **GDS-15 score** |  |  |  |  |  |  |
| Mean (SD) | 1.2 (2.2) | 1.4 (2.1) | 1.3 (2.0) | 0.6346 | 0.6377 | 0.9073 |
| **MDS-UPDRS Part 1 apathy score** |  |  |  |  |  |  |
| % with score ≥1 | 7 (4%)<sup>**</sup> | 8 (8%) | 26 (11%) | 0.2461 | 0.0162<sup>****</sup> | 0.3325 |
| **Meets cognitive criteria for stage 2, n (%)** |  |  |  |  |  |  |
| Item 1.1 = 1 BUT MoCA ≥ 25 | 14 (9%)<sup>***</sup> | 17 (17%) | 37 (16%) | 0.0564 | 0.0461<sup>****</sup> | 0.9045 |
| **Meets motor criteria for stage 2, n (%)** |  |  |  |  |  |  |
| Subthreshold parkinsonism OR PD meds | 5 (3%) | 9 (9%) | 136 (60%) | 0.0461<sup>****</sup> | <.0001<sup>****</sup> | <.0001<sup>****</sup> |
| **Meets non-motor criteria for stage 2, n (%)** |  |  |  |  |  |  |
| Hyposmia OR iRBD | 18 (11%) | 101 (100%) | 227 (100%) | N/A | N/A | N/A |
| **Non-motor subgroup, n (%)** |  |  |  |  |  |  |
| Hyposmia AND iRBD | 0 (0%) | 43 (43%) | 48 (21%) | N/A | N/A | <.0001<sup>****</sup> |
| Hyposmia only | 18 (100%)<sup>***</sup> | 58 (57%) | 179 (79%) |  |  |  |
| **Cohort/Subgroup at Enrollment, n(%)** |  |  |  |  |  |  |
| HC | 158 (100%) | 9 (9%) | 1 (<1%) | N/A | N/A | <.0001<sup>****</sup> |
| Sporadic PD | 0 (0%) | 0 (0%) | 116 (51%) |  |  |  |
| Genetic PD | 0 (0%) | 0 (0%) | 1 (<1%) |  |  |  |
| iRBD | 0 (0%) | 43 (43%) | 48 (21%) |  |  |  |
| Hyposmia | 0 (0%) | 44 (44%) | 58 (26%) |  |  |  |
| Genetic NMC | 0 (0%) | 5 (5%) | 2 (1%) |  |  |  |
| SWEDD | 0 (0%) | 0 (0%) | 1 (<1%) |  |  |  |

\* Robust Healthy Control (HC) Criteria: (a) not S+ (b) MoCA=27 at BL (c) MoCA=26 at 1Y and (d) no more than a 2-point drop in MoCA between BL and 1Y.
\*\* 3 Stage 2B participants had missing education information and 1 HC participant had a missing MDS-UPDRS Part 1 apathy score.
\*\*\* There was 1 HC participant who was not evaluable for cognitive and non-motor criteria due to missing data; there was 1 HC participant who met cognitive criteria for stage 3 i.e., Item 1.1=2 and MoCA ≥ 25.
\*\*\*\* Significance defined at P < 0.05.

### Cognitive performance in Stage 2 NSD with hyposmia

For cognition, MoCA scores were slightly lower for Stage 2A and 2B compared with the HC subgroup, which is not surprising given that MoCA score< 27 was an exclusion criteria for the HC subgroup (**Table 2**). However, the mean MoCA scores for all three groups exceeded the cut-off score (score ≥26).

**Table 2:**
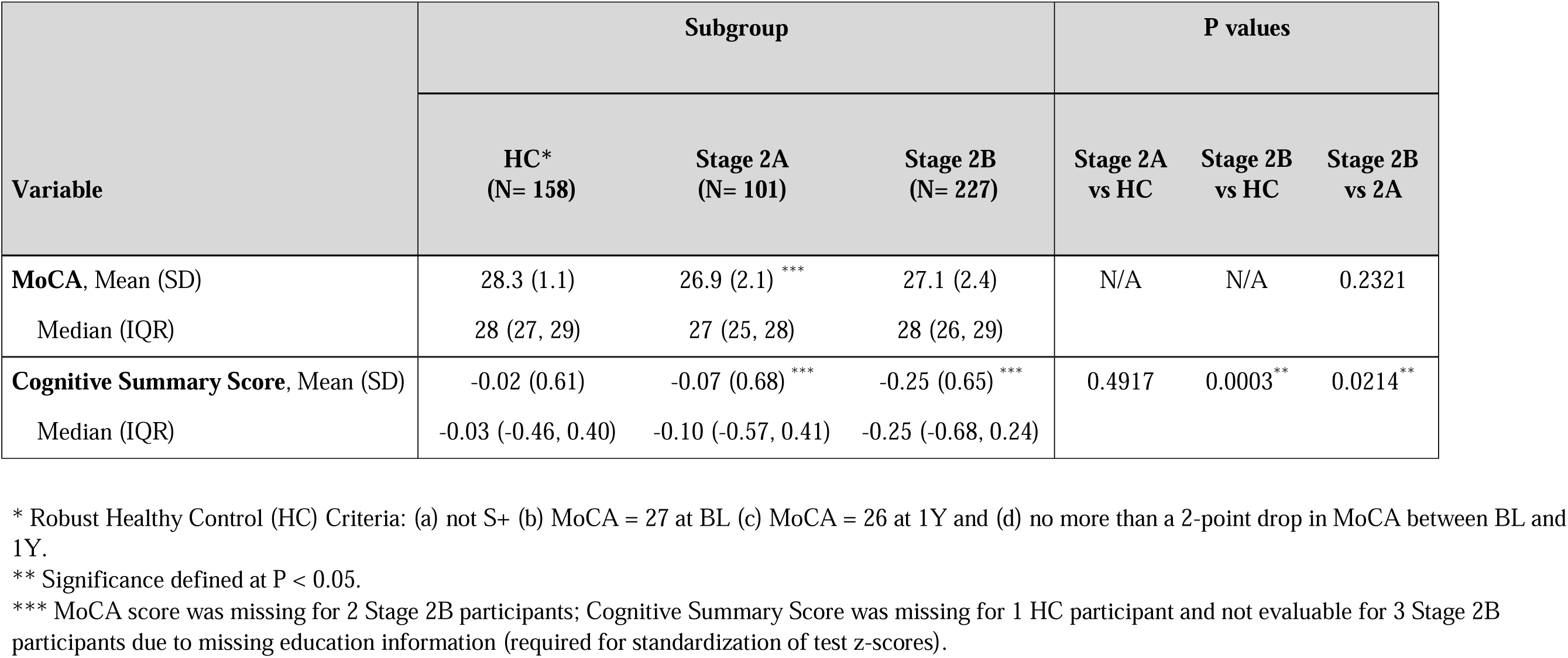
Global cognitive scores by subgroup.

For the CSS, Stage 2A scored numerically worse (z-score mean difference =0.05, p-value NS; effect size=0.09) and Stage 2B scored statistically worse (z-score mean difference =0.23, p-value <0.05; effect size=0.38) than the HC subgroup. Stage 2B scored statistically worse than Stage 2A (z-score mean difference =0.18, p-value <0.05).

### Cognitive performance in Stage 2 NSD with hyposmia by comorbid iRBD

Considering all of Stage 2 NSD as a single group, participants with hyposmia only (i.e., no iRBD) had non-statistically significant worse cognitive performance than the HC subgroup (z-score mean difference =0.12, p-value =NS, effect size =0.18), and having superimposed iRBD nearly doubled the magnitude of cognitive deficits (z-score mean difference for hyposmia + iRBD vs. hyposmia alone =0.24, p-value <0.05, effect size =0.36) (**Table 3-top**).

**Table 3:** Cognitive scores by iRBD status for NSD Stage 2 compared to SHC.

| Variable | HC<br>(N= 158) | Overall Stage 2 |  | P values |  |  |
| --- | --- | --- | --- | --- | --- | --- |
|  |  | Hyposmia Only<br>(N = 237) | Hyposmia + iRBD<br>(N = 91) | Hyposmia Only<br>vs HC | Hyposmia + iRBD<br>vs HC | Hyposmia + iRBD<br>vs Hyposmia Only |
| **Cognitive summary score** |  |  |  | 0.0797 | <.0001 ** | 0.0036 ** |
| Mean (SD) | -0.02 (0.61) | -0.13 (0.66) * | -0.37 (0.67) |  |  |  |
| Median (IQR) | -0.03 (-0.46, 0.40) | -0.12 (-0.63, 0.34) | -0.38 (-0.79, 0.05) |  |  |  |
|  |  | **Stage 2A** |  |  |  |  |
|  |  | **Hyposmia Only**<br>**(N = 58)** | **Hyposmia + iRBD**<br>**(N = 43)** |  |  |  |
| **Cognitive summary score** |  |  |  | 0.3838 | 0.0193 ** | 0.0139 ** |
| Mean (SD) | -0.02 (0.61) | 0.07 (0.71) | -0.26 (0.60) |  |  |  |
| Median (IQR) | -0.03 (-0.46, 0.40) | 0.15 (-0.54, 0.53) | -0.33 (-0.65, 0.03) |  |  |  |
|  |  | **Stage 2B** |  |  |  |  |
|  |  | **Hyposmia Only**<br>**(N = 179)** | **Hyposmia + iRBD**<br>**(N = 48)** |  |  |  |
| **Cognitive summary score** |  |  |  | 0.0078 ** | <.0001 ** | 0.0116 ** |
| Mean (SD) | -0.02 (0.61) | -0.20 (0.63) * | -0.47 (0.72) |  |  |  |
| Median (IQR) | -0.03 (-0.46, 0.40) | -0.21 (-0.64, 0.26) | -0.46 (-0.86, 0.06) |  |  |  |

\* - There were 3 Stage 2B participants in the Hyposmia Only group who had missing Cognitive Summary Scores.
\*\* - Statistically significant at $p < 0.05$ .

Examining NSD Stage 2A participants (i.e., without dopaminergic neuronal dysfunction), those with hyposmia only had normal cognition, but the addition of comorbid iRBD led to a statistically significant increase in cognitive deficit (z-score mean difference for hyposmia + iRBD vs. hyposmia alone =0.33, p-value <0.05, effect size =0.50) (**Table 3-middle; Figure**).

**Figure.**
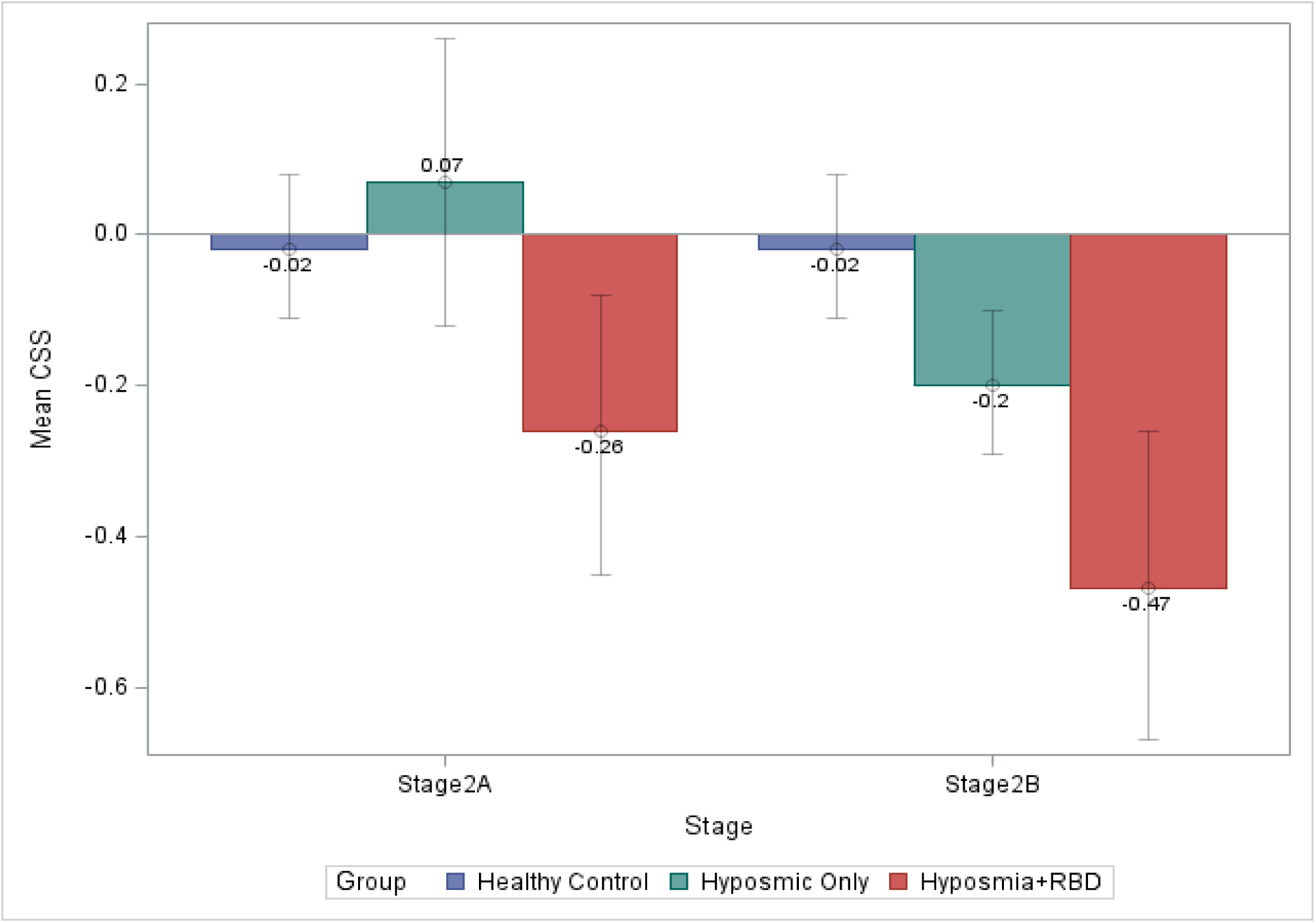
Mean CSS across subgroups in Stage 2A and 2B participants.

In NSD Stage 2B participants (i.e., with dopaminergic neuronal dysfunction), those with hyposmia alone now demonstrated significant cognitive deficits (z-score mean difference= 0.18, p-value < 0.05, effect size=0.29), and superimposed iRBD had an additional statistically significant effect (z-score mean difference =0.27, p-value < 0.05, effect size =0.41) (**Table 3-bottom; Figure**).

## DISCUSSION

Classifying hyposmics with prodromal or *de novo*, untreated PD as early-stage neuronal α-synuclein disease, we found that presence of neuronal α-synuclein disease alone, without concomitant dopaminergic dysfunction, is not associated with cognitive deficits, unless comorbid iRBD is present. On the other hand, once dopaminergic dysfunction is present, hyposmia in and of itself is associated with cognitive changes, and superimposed iRBD continues to have an additive effect. The findings suggest that presence of abnormal synuclein biology in hyposmia does not appear to be associated with cognitive changes until other features emerge, such as dopamine deficiency and iRBD.

Previous research has found that persons in the general population with hyposmia plus dopaminergic system dysfunction (i.e., DAT scan deficit) have detectable cognitive deficits compared with persons without these two characteristics(7, 11). Our results confirm these findings and extend them to participants with diagnosed early-stage NSD, classified on the basis of having a positive α-synuclein SAA test, but with only subtle clinical symptoms and no functional impairment. Interestingly, without dopaminergic neuronal dysfunction cognitive performance was normal in persons with NSD and hyposmia only, suggesting that synucleinopathy at this stage may not be widespread or targeted enough to lead to changes in cognition. Under this hypothesis, cognitive deficits in hyposmics emerge only after dopaminergic dysfunction also occurs, which could reflect either that more widespread brain synuclein disease has developed in parallel or that the dopamine system has a direct role in cognitive decline in NSD, as previously demonstrated in PD using PPMI data(25).

Numerous studies have reported cognitive deficits in patients with iRBD(26), and that cognitive deficits in this disorder predict conversion to dementia with Lewy bodies (DLB) rather than PD(27, 28). Again, we confirmed that the presence of comorbid iRBD in early-stage NSD with hyposmia is associated with clear cognitive changes. Additionally, in contrast to the findings in hyposmia alone, iRBD participants did not require that a dopaminergic deficit be present to demonstrate cognitive decline, suggesting either that iRBD is associated with more diffuse synuclein disease even in early disease, co-occurs with significant non-dopaminergic neurotransmitter deficits that contribute to cognitive impairment, or commonly co-occurs with other disease pathology impacting cognition (e.g., Alzheimer’s disease, which is known to be common in Lewy body dementias(29)). So, as previously reported, although dopaminergic deficits in iRBD predict faster conversion to alpha-synucleinopathy(30), they don’t appear essential to the cognitive decline that occurs in this population.

As alluded to above, the neurobiological underpinnings of cognitive changes in prodromal Lewy body disorders, called here early-stage NSD, appear complex. Overall, just the presence of synuclein disease and prodromal clinical features, in this case hyposmia and to some extent comorbid iRBD, is associated with detectable cognitive differences, suggesting a possible role for synuclein in cognitive performance, even in early disease. The addition of dopaminergic deficiency has an additive effect on cognitive impairment, already reported in early PD for fronto-striatal executive impairments(31). Additionally, the association with cognitive dysfunction appears more profound for iRBD compared with hyposmia alone, suggesting that the cognitive changes seen in these two disorders may have different neurobiological underpinnings. This is consistent with iRBD having a more malignant cognitive course than hyposmia, with more frequent conversion to DLB in iRBD(32) than is seen with hyposmia, which progresses primarily to PD(33, 34). These differences are also included the “brain-first versus gut-first” model of Lewy body disorders(35), with the former including hyposmia as an initial symptom and only limited cognitive decline early on, and the latter including iRBD as an initial disorder with significant early cognitive decline(36).

Strengths of the study include a focus on two prodromal conditions for Lewy body disorders (i.e., hyposmia and iRBD), a large sample size, inclusion of both biological anchors for NSD-ISS (i.e., CSF α-synuclein SAA and DaTscan), and use of a cognitive summary score derived from a detailed cognitive battery, normed using an internal, robust HC group, and that controlled for effects of age, sex and education. Regarding the last point, it’s interesting to note that although the MoCA score was lower and the percentage meeting cognitive criteria for Stage 2 was higher in Stage 2A than in Stage 2B, the CSS score was lower in Stage 2B, suggesting a benefit in utilizing a more sensitive cognitive measure. Limitations include not having an adequate sample size of iRBD participants without hyposmia as a comparator group, allowing Stage 2 NSD to have been enrolled in the PPMI study as part of different clinical cohorts, and only having a dichotomous outcome for SAA status. Regarding the latter point, there is preliminary evidence that a continuous measure of neuronal α-synuclein SAA (i.e., kinetic parameters) is associated with cognitive performance in PD(37), so future research can examine if there is a correlation between this measure and cognitive performance in Stage 2 NSD. Additionally, presence of comorbid AD pathology and other reported biological contributors to cognitive decline in PD dementia or DLB can be examined to see if they help explain the greater cognitive deficits seen in participants with comorbid iRBD.

The findings of detectable cognitive differences in early-stage NSD with hyposmia, particularly in those with dopaminergic deficiency or comorbid iRBD, support the inclusion in NSD-ISS of a cognitive track for symptom progression and functional impairment at all disease stages. The results highlight the importance of carefully assessing cognition, with more than a screening instrument, in longitudinal observational studies and in randomized clinical trials of early-stage neuronal alpha-synuclein disease.

## AUTHOR CONTRIBUTIONS

DW takes responsibility for the integrity of the work as a whole, from inception to published article. All other authors made substantial contributions to conception or design of the work, or the acquisition, analysis, or interpretation of data for the work; and drafting of the work or reviewing it critically for important intellectual content; and final approval of the version to be published; and agreement to be accountable for all aspects of the work in ensuring that questions related to the accuracy or integrity of any part of the work are appropriately investigated and resolved.

## PPMI STUDY TEAMS/CORES/COLLABORATORS FOR PUBLICATIONS

### Executive Steering Committee

Kenneth Marek, MD^1^ (Principal Investigator); Caroline Tanner, MD, PhD^8^; Tanya Simuni, MD^3^; Andrew Siderowf, MD, MSCE^11^; Douglas Galasko, MD^26^; Lana Chahine, MD^38^; Christopher Coffey, PhD^4^; Kalpana Merchant, PhD^58^; Kathleen Poston, MD^37^; Roseanne Dobkin, PhD^40^; Tatiana Foroud, PhD^14^; Brit Mollenhauer, MD^7^; Dan Weintraub, MD^11^; Ethan Brown, MD^8^; Karl Kieburtz, MD, MPH^22^; Mark Frasier, PhD^5^; Todd Sherer, PhD^5^; Sohini Chowdhury, MA^5^; Roy Alcalay, MD^45^ and Aleksandar Videnovic, MD^44^

### Steering Committee

Duygu Tosun-Turgut, PhD^8^; Werner Poewe, MD^6^; Susan Bressman, MD^13^; Jan Hammer^14^; Raymond James, RN^21^; Ekemini Riley, PhD^39^; John Seibyl, MD^1^; Leslie Shaw, PhD^11^; David Standaert, MD, PhD^17^; Sneha Mantri, MD, MS^59^; Nabila Dahodwala, MD^11^; Michael Schwarzschild^44^; Connie Marras^42^; Hubert Fernandez, MD^24^; Ira Shoulson, MD^22^; Helen Rowbotham^2^; Paola Casalin^10^ and Claudia Trenkwalder, MD^7^

### Michael J. Fox Foundation (Sponsor)

Todd Sherer, PhD; Sohini Chowdhury, MA; Mark Frasier, PhD; Jamie Eberling, PhD; Katie Kopil, PhD; Alyssa O’Grady; Maggie McGuire Kuhl; Leslie Kirsch, EdD and Tawny Willson, MBS

### Study Cores, Committees and Related Studies

*Project Management Core:* Emily Flagg, BA^1^

*Site Management Core:* Tanya Simuni, MD^3^; Bridget McMahon, BS^1^

Strategy and Technical Operations: Craig Stanley, PhD^1^; Kim Fabrizio, BA^1^

*Data Management Core:* Dixie Ecklund, MBA, MSN^4^; Trevis Huff, BSE^4^

*Screening Core:* Tatiana Foroud, PhD^14^; Laura Heathers, BA^14^; Christopher Hobbick, BSCE^14^; Gena Antonopoulos, BSN^14^

*Imaging Core:* John Seibyl, MD^1^; Kathleen Poston, MD^37^

*Statistics Core*: Christopher Coffey, PhD^4^; Chelsea Caspell-Garcia, MS^4^; Michael Brumm, MS^4^

*Bioinformatics Core*: Arthur Toga, PhD^9^; Karen Crawford, MLIS^9^

*Biorepository Core:* Tatiana Foroud, PhD^14^; Jan Hamer, BS^14^

*Biologics Working Group:* Brit Mollenhauer^7^; Doug Galasko^26^;

*Biospecimen Review Committee*: Kalpana Merchant^58^

*Genetics Core:* Andrew Singleton, PhD^12^

*Pathology Core:* Tatiana Foroud, PhD^14^; Thomas Montine, MD, PhD^37^

*Found:* Caroline Tanner, MD PhD^8^

*PPMI Online:* Carlie Tanner, MD PhD^8^; Ethan Brown, MD^8^; Lana Chahine, MD^38^; Roseann Dobkin, PhD^40^; Monica Korell, MPH^8^

### Site Investigators

Charles Adler, PhD^48^; Roy Alcalay, MD^34^; Amy Amara, PhD^49^; Paolo Barone, PhD^29^; Bastiaan Bloem, PhD^57^ Susan Bressman, MD^15^; Kathrin Brockmann, MD^25^; Norbert Brüggemann, MD^56^; Lana Chahine, MD^38^; Kelvin Chou, MD^41^; Nabila Dahodwala, MD^11^; Alberto Espay, MD^31^; Stewart Factor, DO^15^; Hubert Fernandez, MD^24^; Michelle Fullard, MD^49^; Douglas Galasko, MD^26^; Robert Hauser, MD^18^; Penelope Hogarth, MD^16^; Shu-Ching Hu, PhD^20^; Michele Hu, PhD^55^; Stuart Isaacson, MD^30^; Christine Klein, MD^56^; Rejko Krueger, MD^2^; Mark Lew, MD^46^; Zoltan Mari, MD^53^; Connie Marras, PhD^42^; Maria Jose Martí, PhD^32^; Nikolaus McFarland, PhD^51^; Tiago Mestre, PhD^43^; Brit Mollenhauer, MD^7^; Emile Moukheiber, MD^27^; Alastair Noyce, PhD^60^; Wolfgang Oertel, PhD^61^; Njideka Okubadejo, MD^62^; Sarah O’Shea, MD^36^; Rajesh Pahwa, MD^45^; Nicola Pavese, PhD^54^; Werner Poewe, MD^6^; Ron Postuma, MD^52^; Giulietta Riboldi, MD^50^; Lauren Ruffrage, MS^17^; Javier Ruiz Martinez, PhD^33^; David Russell, PhD^1^; Marie H Saint-Hilaire, MD^21^; Neil Santos, BS^48^; Wesley Schlett^44^; Ruth Schneider, MD^22^; Holly Shill, MD^47^; David Shprecher, DO^23^; Tanya Simuni, MD^3^; David Standaert, PhD^17^; Leonidas Stefanis, PhD^35^; Yen Tai, PhD^28^; Caroline Tanner, PhD^8^; Arjun Tarakad, MD^19^; Eduardo Tolosa PhD^32^ and Aleksandar Videnovic, MD^44^

### Coordinators

Susan Ainscough, BA^29^; Courtney Blair, MA^17^; Erica Botting^18^; Isabella Chung, BS^53^; Kelly Clark^23^; Ioana Croitoru^33^; Kelly DeLano, MS^31^; Iris Egner, PhD^6^; Fahrial Esha, BS^50^; May Eshel, MSc^34^; Frank Ferrari, BS^41^; Victoria Kate Foster^54^; Alicia Garrido, MD^32^; Madita Grümmer^56^; Bethzaida Herrera^47^; Ella Hilt^25^; Chloe Huntzinger, BA^49^; Raymond James, BS^21^; Farah Kausar, PhD^8^; Christos Koros, MD, PhD^35^; Yara Krasowski, MSc^57^; Dustin Le, BS^16^; Ying Liu, MD^49^; Taina M. Marques, PhD^2^; Helen Mejia Santana, MA^36^; Sherri Mosovsky, MPH^38^; Jennifer Mule, BS^24^; Philip Ng, BS^42^; Lauren O’Brien^45^; Abiola Ogunleye, PGDip^28^; Oluwadamilola Ojo, MD^62^; Obi Onyinanya, BS^27^; Lisbeth Pennente, BA^30^; Romina Perrotti^52^; Michael Pileggi, MS^52^; Ashwini Ramachandran, MSc^11^; Deborah Raymond, MS^13^; Jamil Razzaque, MS^55^; Shawna Reddie, BA^43^; Kori Ribb, BSN,^27^; Kyle Rizer, BA^51^; Janelle Rodriguez, BS^26^; Stephanie Roman, HS^1^; Clarissa Sanchez, MPH^19^; Cristina Simonet, PhD^28^; Anisha Singh, BS^22^; Elisabeth Sittig, RN^61^; Barbara Sommerfeld MSN^15^; Angela Stovall, BS^41^; Bobbie Stubbeman, BS^31^; Alejandra Valenzuela, BS^46^; Catherine Wandell, BS^20^; Diana Willeke^7^; Karen Williams, BA^3^ and Dilinuer Wubuli, MB^42^

### Partners Scientific Advisory Board (Acknowledgement)

Funding: PPMI – a public-private partnership – is funded by the Michael J. Fox Foundation for Parkinson’s Research and funding partners, including 4D Pharma, Abbvie, AcureX, Allergan, Amathus Therapeutics, Aligning Science Across Parkinson’s, AskBio, Avid Radiopharmaceuticals, BIAL, BioArctic, Biogen, Biohaven, BioLegend, BlueRock Therapeutics, Bristol-Myers Squibb, Calico Labs, Capsida Biotherapeutics, Celgene, Cerevel Therapeutics, Coave Therapeutics, DaCapo Brainscience, Denali, Edmond J. Safra Foundation, Eli Lilly, Gain Therapeutics, GE HealthCare, Genentech, GSK, Golub Capital, Handl Therapeutics, Insitro, Jazz Pharmaceuticals, Johnson & Johnson Innovative Medicine, Lundbeck, Merck, Meso Scale Discovery, Mission Therapeutics, Neurocrine Biosciences, Neuron23, Neuropore, Pfizer, Piramal, Prevail Therapeutics, Roche, Sanofi, Servier, Sun Pharma Advanced Research Company, Takeda, Teva, UCB, Vanqua Bio, Verily, Voyager Therapeutics, the Weston Family Foundation and Yumanity Therapeutics.

1. Institute for Neurodegenerative Disorders, New Haven, CT
2. University of Luxembourg, Luxembourg
3. Northwestern University, Chicago, IL
4. University of Iowa, Iowa City, IA
5. The Michael J. Fox Foundation for Parkinson’s Research, New York, NY
6. Innsbruck Medical University, Innsbruck, Austria
7. Paracelsus-Elena Klinik, Kassel, Germany
8. University of California, San Francisco, CA
9. Laboratory of Neuroimaging (LONI), University of Southern California
10. BioRep, Milan, Italy
11. University of Pennsylvania, Philadelphia, PA
12. National Institute on Aging, NIH, Bethesda, MD
13. Mount Sinai Beth Israel, New York, NY
14. Indiana University, Indianapolis, IN
15. Emory University of Medicine, Atlanta, GA
16. Oregon Health and Science University, Portland, OR
17. University of Alabama at Birmingham, Birmingham, AL
18. University of South Florida, Tampa, FL
19. Baylor College of Medicine, Houston, TX
20. University of Washington, Seattle, WA
21. Boston University, Boston, MA
22. University of Rochester, Rochester, NY
23. Banner Research Institute, Sun City, AZ
24. Cleveland Clinic, Cleveland, OH
25. University of Tübingen, Tübingen, Germany
26. University of California, San Diego, CA
27. Johns Hopkins University, Baltimore, MD
28. Imperial College of London, London, UK
29. University of Salerno, Salerno, Italy
30. Parkinson’s Disease and Movement Disorders Center, Boca Raton, FL
31. University of Cincinnati, Cincinnati, OH
32. Hospital Clinic of Barcelona, Barcelona, Spain
33. Hospital Universitario Donostia, San Sebastian, Spain
34. Tel Aviv Sourasky Medical Center, Tel Aviv, Israel
35. National and Kapodistrian University of Athens, Athens, Greece
36. Columbia University Irving Medical Center, New York, NY
37. Stanford University, Stanford, CA
38. University of Pittsburgh, Pittsburgh, PA
39. Center for Strategy Philanthropy at Milken Institute, Washington D.C.
40. Rutgers University, Robert Wood Johnson Medical School, New Brunswick, New Jersey
41. University of Michigan, Ann Arbor, MI
42. Toronto Western Hospital, Toronto, Canada
43. The Ottawa Hospital, Ottawa, Canada
44. Massachusetts General Hospital, Boston, MA
45. University of Kansas Medical Center, Kansas City, KS
46. University of Southern California, Los Angeles, CA
47. Barrow Neurological Institute, Phoenix, AZ
48. Mayo Clinic Arizona, Scottsdale, AZ
49. University of Colorado, Aurora, CO
50. NYU Langone Medical Center, New York, NY
51. University of Florida, Gainesville, FL
52. Montreal Neurological Institute and Hospital/McGill, Montreal, QC, Canada
53. Cleveland Clinic-Las Vegas Lou Ruvo Center for Brain Health, Las Vegas, NV
54. Clinical Ageing Research Unit, Newcastle, UK
55. John Radcliffe Hospital Oxford and Oxford University, Oxford, UK
56. Universität Lübeck, Luebeck, Germany
57. Radboud University, Nijmegen, Netherlands
58. TransThera Consulting
59. Duke University, Durham, NC
60. Wolfson Institute of Population Health, Queen Mary University of London, UK
61. Philipps-University Marburg, Germany
62. University of Lagos, Nigeria

